# The Progressive Aphasia Communication Toolkit (PACT): A Strengths-Based Approach to Multidomain Evaluation for Intervention

**DOI:** 10.64898/2025.11.25.25340904

**Authors:** Jeanne Gallée, Jade Cartwright, Maya L Henry, Aimee R Mooney, Brielle C Stark, Anna Volkmer, Aimee Dietz, Connie Nakano, Petronilla Battista, Ashleigh Beales, Bárbara Costa Beber, Inês Cadório, Megan Caldwell, Katharine Davies, Zoe Ezzes, Mirjam Gauch, Tracey Graney, Simon Grobler, Katarina L Haley, Alessa Hausmann, Elena Herrera, Honey I. Hubbard, Regina Jokel, Leslie Kot, Mara Lowe, Ellen McGowan, Lotte Meteyard, Núria Montagut, Iris Nowenstein, Margaret Pozzebon, Richard Talbot, Cathy Taylor-Rubin, Ffion Walker, Ingvild Elisabeth Winsnes, Rob J Fredericksen, Kimiko Domoto-Reilly, Paul K Crane

**Affiliations:** Department of Medicine, University of Washington, Seattle; Alzheimer’s Disease Research Center, University of Washington, Seattle; School of Health Sciences, University of Tasmania, Hobart, Australia; Department of Speech, Language, and Hearing Sciences, University of Texas-Austin, Austin, TX, United States of America; Department of Neurology, Oregon Health & Science University, Portland, OR, United States of America; Department of Speech, Language, and Hearing Sciences, Indiana University, Bloomington, IN, United States of America; Division of Psychology and Language Sciences, University College London, London; Department of Communication Sciences and Disorders, University of South Florida, Tampa, FL, USA; Istituti Clinici Scientifici Maugeri IRCCS, Laboratory of Neuropsychology, Institute of Bari, Bari, Italy; Speech Rehab Centre, Perth, Australia; Department of Speech, Language and Hearing Sciences, Universidade Federal de Ciências da Saúde de Porto Alegre (UFCSPA), Porto Alegre, Rio Grande do Sul, Brazil; RISE-Health, Escola Superior de Saúde Fernando Pessoa, Fundação Ensino e Cultura Fernando Pessoa, Porto, Portugal; Instituto de Investigação, Inovação e Desenvolvimento (FP-I3ID), Unidade de Investigação de Ciências Biomédicas e da Saúde (FP-BHS), Universidade Fernando Pessoa, Porto, Portugal; Escola Superior de Saúde Fernando Pessoa (ESS-FP), Porto, Portugal; Department of Speech and Hearing Sciences, University of Washington, Seattle, WA, USA; School of Audiology and Speech Sciences, The University of British Columbia, Vancouver, Canada; School of Speech, Language, and Hearing Sciences, San Diego State University, San Diego, CA, USA; Department of Cognitive Science, University of California, San Diego, San Diego, CA, USA; Department of Psychiatry and Psychotherapy, University Medical Center of the Johannes Gutenberg University Mainz, 55131 Mainz, Germany; Tracey Graney Speech Pathology, Hobart, Tasmania, Australia; National Hospital for Neurology and Neurosurgery, London, UK; Division of Speech and Hearing Sciences, Department of Health Sciences, School of Medicine, University of North Carolina, Chapel Hill, NC, USA; Department of Psychology, University of Geneva, Geneva, Switzerland; Department of Clinical Neurosciences, Lausanne University Hospital and University of Lausanne, Lausanne, Switzerland; Department of Psychology, University of Oviedo, Oviedo, Spain; Department of Speech and Hearing Sciences, University of New Mexico, Albuquerque, NM, USA; Rotman Research Institute, Baycrest Academy for Research and Education, Toronto, Canada; Department of Speech-Language Pathology, Temerty Faculty of Medicine, University of Toronto, Toronto, Canada; Baycrest Health Sciences, Toronto, Canada; Department of Linguistics and Communication Disorders, CUNY Queens College, New York, NY, USA; Pennine Care NHS Foundation Trust, Manchester, UK; Clinical Language Sciences, University of Reading, UK; Alzheimer’s Disease and other Cognitive Disorders Unit. Hospital Clínic de Barcelona, Barcelona, Spain; Fundació de Recerca Clínic Barcelona-IDIBAPS, Barcelona, Spain; Department of Icelandic, University of Iceland, Reykjavík, Iceland; Speech-Language Pathology Unit, National University Hospital, Reykjavík, Iceland; Age Right Speech Pathology, Melbourne, Australia; Taylor-Rubin Speech Language Pathology, Sydney, Australia; HealthAbility, Melbourne, Australia; Department of Linguistics and Scandinavian Studies, University of Oslo, Oslo, Norway; Department of Allergy and Infectious Diseases, University of Washington, Seattle, WA, USA; Department of Neurology, University of Washington, Seattle, WA, USA

**Keywords:** primary progressive aphasia, therapeutic assessment, strengths-based approach, person-centered care, speech-language pathology

## Abstract

**INTRODUCTION:** Tools to document communicative ability in people living with primary progressive aphasia (PwPPA) are limited. This work describes the development of a strengths-based and ecologically-valid instrument—the *Progressive Aphasia Communication Toolkit* (PACT).

**METHODS:** This work consisted of five experiments: two to develop (Experiments 1 and 2) and three to pilot (Experiments 3-5) a novel instrument for PPA. Ninety-five individuals worldwide contributed to this work: 80 researchers and clinicians, 9 PwPPA, and 6 care partners.

**RESULTS:** Experiments 1-2 culminated in an instrument comprising four scales that capture quantitative and qualitative feedback. Experiments 3-5 resulted in structural refinement and digitization of the tool, revealed PwPPA and care partner preference for the PACT over traditional neuropsychological evaluation, and demonstrated strong inter-rater agreement for general measurability (91%) and strength ratings (85%).

**DISCUSSION:** Current findings indicate that the PACT provides a holistic profile of communication strengths for PwPPA and can guide clinicians in developing functional therapeutic targets.

## 1 BACKGROUND

Primary progressive aphasia (PPA) is increasingly recognized as a condition that progressively impacts both communication and life participation.^1–5^ Current efforts to characterize communication in people living with PPA (PwPPA) are constrained by assessment tools that prioritize diagnostic classification over documenting communicative competence.^6^ Established language assessments used in the care of PwPPA are heavily impairment-focused and provide little insight into conserved communication abilities or compensatory strategy use,^7,8^ critically limiting their ecological validity^1,9^ clinical relevance for intervention planning.^1,4,6,7,8^ Additionally, most existing aphasia assessment tools were developed for sequelae of stroke and thus lack sensitivity to the unique challenges of PPA.^1,6,7^ Despite advances in characterizing the symptomatic profiles of PPA subtypes,^10–17^ the functional relevance of these behaviors for overall communicative ability remains insufficiently documented and interpreted.^1,7,18^ These shortcomings have profound effects on clinical decision-making, hindering person-centered care, obscuring client autonomy, and delaying the development of proactive intervention plans for this population.^1,7,18^ Consequently, there is an urgent need for instruments that capture communication in real-world contexts and support a strengths-based approach to assessment and treatment.^1–9,18–20^

Beyond sensitivity and ecological validity, an instrument designed for PPA must minimize burden.^4,6^ Progressive conditions like PPA require regular, frequent reassessment to monitor current status and symptom evolution. Repeated testing can be burdensome, strenuous, and feel arbitrary.^4,6,7^ Assessment outcomes may also not be relayed to the individual who was tested, nor offer practical insight beyond diagnostic characterization.^4,6,7^ A recent survey of speech-language pathologists (SLPs) working with PwPPA worldwide identified a need for tools specifically designed for PPA.^7^ In line with the R.A.I.S.E. Assessment Framework for PPA,^4^ assessment ideally integrates clinician, client, and care partner feedback^1,7,9,21^ to prioritize the client and ensure reciprocity of benefit. Finally, the ideal instrument is adaptable to evolving client needs and clinical presentation (e.g., by allowing for text-based, non-verbal, or multimodal communication^2,4,6^). Therefore, the field requires a validated and reliable measure that (1) is specifically developed for PPA, (2) minimizes patient burden, and (3) captures communication ability with ecological and clinical relevance for both research and practice.

We therefore set out to develop a person-centered instrument for PPA through a multitiered process involving SLPs, clinical researchers, PwPPA, and care partners. Our aim was to create a tool that leverages conversational output from clinical or research visits to document behavioral strengths across major communication domains, and to allow findings to be easily across institutions and health professions.

## 2 Methods

The methods of this work adhere to the Gallée et al. (2024) protocol^20^ (**Figure 1**). Ethical approval for each of the study arms was obtained from the University of Washington (UW) Human Subjects Division (HSD) on January 3^rd^, 2024 (“Assessment of Communicative Ability in Alzheimer’s Disease and Related Dementias; STUDY00019344).

**Figure 1.**
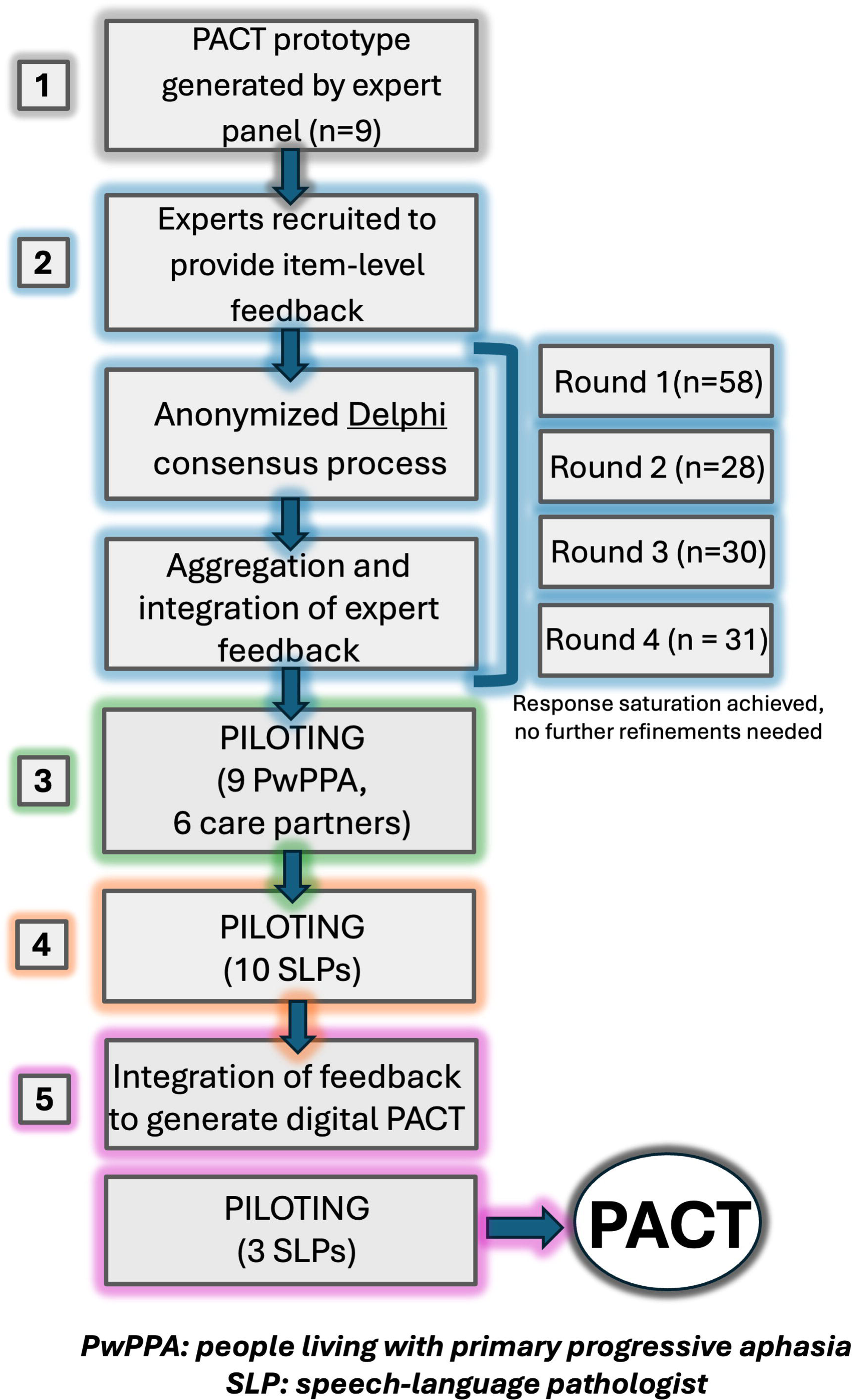
Study the flowchart of the five experiments guiding the PACT development. Experiment 1 (gray) generated the original PACT prototype, as outlined in Gallée et al. (2024). Experiment 2 (light blue) consisted of soliciting and generating feedback through the Delphi consensus process. Experiment 3 (green) piloted the PACT protocol with nine PwPPA and six care partners. Experiment 4 (orange) piloted completing the PACT assessment form with 10 speech-language pathologists (SLPs). Experiment 5 (pink) piloted the updated and digitalized PACT with three additional SLPs.

### 2.1 Experiment 1: Prototype Generation

#### 2.1.1 Participants

Nine clinical researchers participated in the original drafting of the scale, seven of whom are licensed healthcare professionals across fields. See **Supplementary Table 1** for a description of the expert panel that collaboratively built the instrument prototype.

#### 2.1.2 Methods

Authors JG, KDR, RJF, and PKC generated a rating scale and initial inventory of cognitive-linguistic behaviors. Item selection was guided by Kirshner & Guyatt (1985)’s methodological framework for health indices.^22^ Authors JC, MLH, AM, BCS, and AV met with JG across three Zoom meetings to revise the inventory. Further modifications were made over email until each panelist affirmed satisfaction with the preliminary instrument.

#### 2.1.3 Results

Five domains were presented in the instrument prototype: *Speech and Voice* (9 items), *Language* (9 items), *Social-Pragmatics* (11 items), *Discourse* (8 items), and *Cognition* (9 items). See **Supplementary Table 2A** for the complete prototype.

### 2.2 Experiment 2: Expert Consensus

#### 2.2.1 Participants

To develop an instrument with expert feedback, ninety-seven individuals with expertise in PPA and/or functional communication in acquired neurogenic communication disorders were contacted to participate in an electronic Delphi process^23–25^ (see **Table 1** for participant demographics). Participants were recruited via network sampling, beginning with the International SLT/P PPA Network,^26^ and contacted via email. Participants who continued past the first two rounds were invited to co-author the resulting manuscript for this work. Participants could opt out of the study at any time.

**Table 1.**
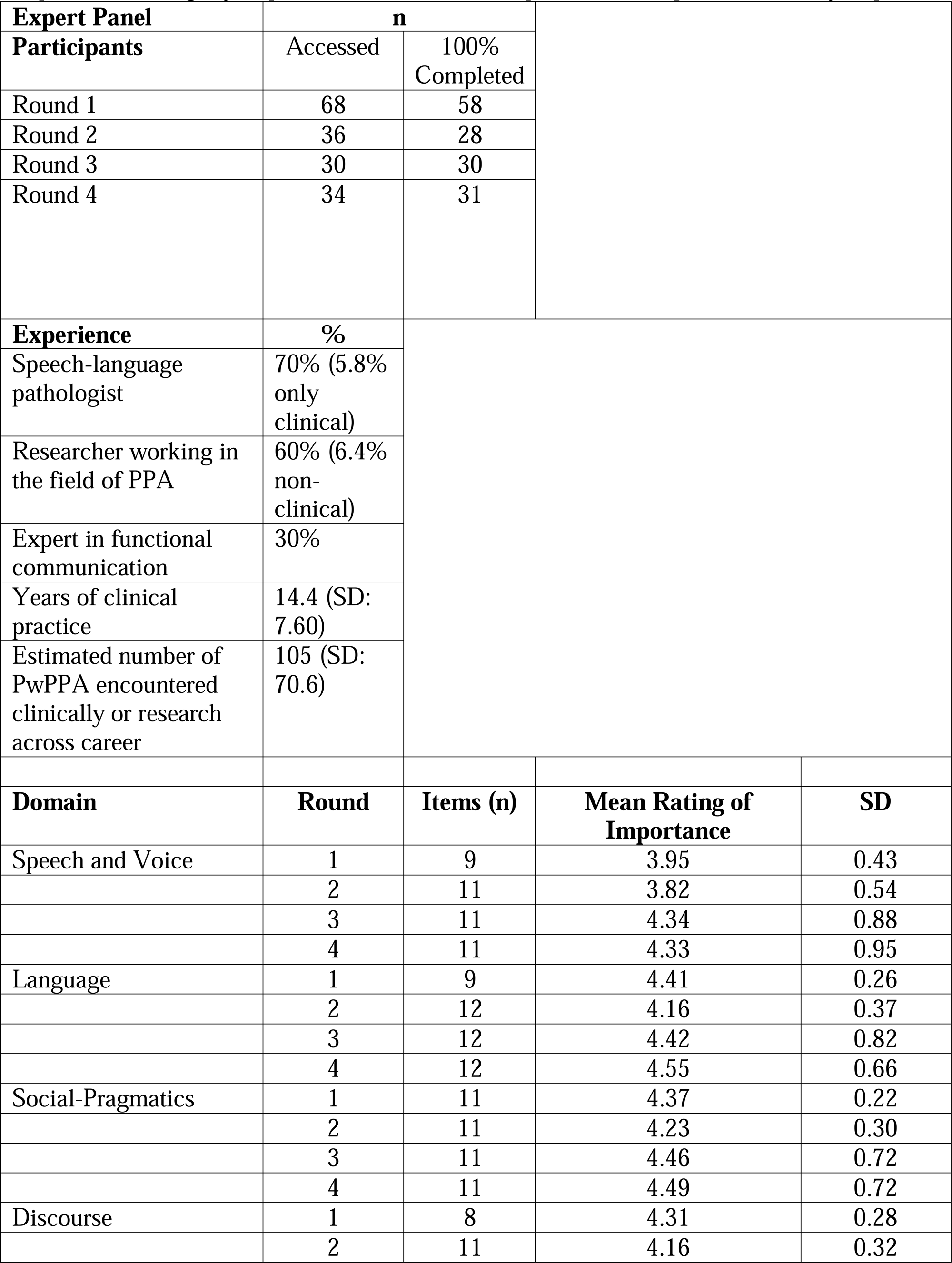
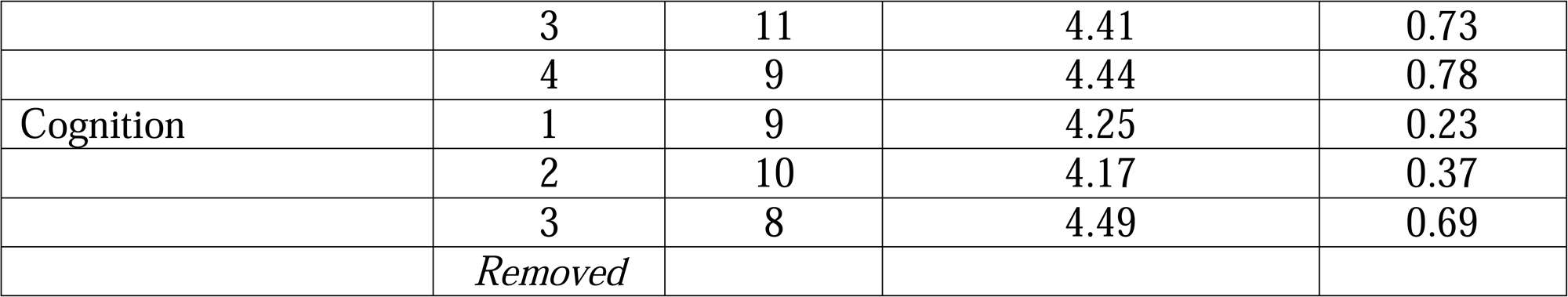
Self-reported experience of Delphi consensus process participants and Delphi consensus outcomes. Participants ranked item importance on a scale from 1-5 (1 = not at all important, 2 = slightly important, 3 = somewhat important, 4 = important, 5 = very important).

#### 2.2.2 Methods

The online survey for the Delphi consensus process was developed by the core study authors (2.1.1) Data were collected anonymously, with IP addresses suppressed, via the Qualtrics^®^ survey platform (Qualtrics, Provo, UT).^27^ Survey respondents could review their responses and skip over any survey items. Each of the four surveys solicited consent to participate and non-identifiable demographic information. Participants were asked to rate the importance of each proposed item (1 = not at all important to 5 = very important) and each overall domain (1 = not at all important to 5 = very important). Survey respondents were provided with figures outlining the intended instrument format to guide their ratings. Open-ended text boxes prompted survey respondents to document section-specific recommendations, identify missing or duplicative content, and provide general feedback on the measurement tool. Following the first round, authors JG and RJF identified, summarized, and integrated respondent feedback by content type (e.g., rank order rationale, rationale for proposed alternative/revised item, etc.) prior to enacting a qualitative process based on the Braun and Clarke (2006)^28^ model of thematic analysis to summarize themes within each content type (e.g., by familiarizing oneself with the data, generating preliminary codes, identifying potential themes, coding the data by themes, and summarizing the outcomes accordingly). Investigators independently summarized themes and subsequently met to discuss and reconcile differences in interpretation and achieve consensus. The reconciled interpretations were then used to iteratively adapt and restructure the measurement prototype (see **Supplementary Table 2B** for summarized feedback). Data collection took place between May 7^th^, 2024, and July 8^th^, 2025.

#### 2.2.3 Results

Sixty-eight participants accessed the online survey, of whom 58 provided complete feedback on at least one Delphi round. Thirty-one participants provided feedback on the last round, where 70% self-identified as SLPs, 60% as researchers focused on PPA, and 30% as experts in the study of functional communication. Respondents reported having an average of 14.4 years of clinical practice (SD: 7.60) and having seen 105 PwPPA in their careers (SD: 70.6). The five domains of the measurement prototype received ratings between “somewhat important” and “very important” from the first Delphi round. Following Round 1, survey respondents received a quantitative and qualitative summary of participant feedback, explanations of the proposed changes, and an opportunity to evaluate the newest prototype.

By Round 3, the instrument was renamed^16^ the *Progressive Aphasia Communication Toolkit* (PACT) to better reflect its aims and structure. *Cognition* was removed based on respondent feedback and panel opinion that a single conversational sample was insufficient to evaluate this domain without additional standardized testing. By Round 4, response saturation was achieved and respondents rated the PACT items as “important” (4) to “very important” (5) for scales of *Speech and Voice* (median: 5.0, range: 1.0-5.0, IQR: 1), *Language* (median: 5.0, range: 2.0-5.0, IQR: 1), *Social-Pragmatics* (median: 5.0, range: 2.0-5.0, IQR: 1) and *Discourse* (median: 5.0, range: 2.0-5.0, IQR: 1; see **Table 1** and **Supplementary Tables 2A and 2B**).

### 2.3 Experiment 3: Piloting the PACT with PwPPA and Care Partners

#### 2.3.1. Participants

To explore the acceptability of the PACT, nine individuals diagnosed with svPPA (n = 1), nfvPPA (n = 2), lvPPA (n = 3), or a mixed presentation (n = 3) were recruited (see **Table 2**). Six care partners of these individuals co-participated. Participants were eligible to participate in the study if they had received a diagnosis of PPA at the University of Washington (UW), were enrolled in the UW Alzheimer’s Disease Research Center (ADRC) Clinical Core, the UW ADRC Research Registry, or the UW Speech and Hearing Clinic, and demonstrated capacity to comply with the experimental protocol. Participants were ineligible to participate if they did not demonstrate capacity to comply with the experimental protocol due to hearing, English proficiency, vision, or cognitive impairments. Similarly, participants who did not consent to being video- and audio-recorded for the PACT protocol were excluded. All participants received a full explanation of the project from study staff. Informed consent was obtained from all participants, in line with the recruitment procedures and experimental protocol approved by the UW IRB. A copy of the signed consent form was provided to all enrollees. Research visits were conducted either at the UW ADRC (n = 3) or during a home visit (n = 6). Participants were paid $150 for their time with an online gift card.

**Table 2.** Participant demographics for the conversational sample aspect of the PACT protocol. Participants for whom the recordings were used to pilot the assessment form are marked by *. “Care partner present” indicates whether the care partner of the client participated in the PACT protocol recording. Conversation length denotes the total time the participant engaged in conversation, beyond the PACT protocol itself.

| Participant | Diagnosis | Sex | Age range (years) | Age range of symptom onset (years) | Education (years) | CDR | Reports subjective decline in memory? | Standardized assessment scores | Care partner present | Conversation length (min:sec) |
| --- | --- | --- | --- | --- | --- | --- | --- | --- | --- | --- |
| 1* | svPPA<sup>1</sup> | M | 66-70 | 51-55 | 20 | 0.5 | No | MoCA<sup>6</sup>: 21/30<br>QAB<sup>7</sup>: moderate | Yes | 14:47 |
| 2* | nfvPPA<sup>2</sup> | F | 81-85 | 71-75 | 14 | 0.5 | No | MoCA: 25/30<br>QAB: mild | No | 14:22 |
| 3 | nfvPPA/CBS<sup>3</sup> | F | 61-65 | 56-60 | 14 | 0 | No | MoCA: 23/30 | Yes | 4:55 |
| 4* | Mixed<sup>4</sup> | F | 56-60 | 51-55 | 16 | 0.5 | No | QAB: severe | Yes | 17:21 |
| 5 | Mixed | M | 76-80 | 71-75 | 21 | 1 | Yes | Declined | Yes | 15:44 |
| 6 | Mixed | M | 76-80 | 71-75 | 16 | 2 | Yes | Declined | Yes | 31:44 |
| 7* | lvPPA<sup>5</sup> | M | 71-75 | 66-70 | 16 | 0.5 | No | QAB: mild | No | 31:36 |
| 8 | lvPPA | F | 76-80 | 76-80 | 18 | 0.5 | Yes | MoCA: 22/30<br>Declined QAB | No | 23:52 |
| 9 | lvPPA | M | 71-75 | 61-65 | 22 | 0.5 | No | QAB: mild | Yes | 24:39 |

<sup>1</sup>svPPA: semantic variant primary progressive aphasia
<sup>2</sup>nfvPPA: nonfluent variant primary progressive aphasia
<sup>3</sup>CBS: corticobasal syndrome
<sup>4</sup>Mixed: mixed presentation of primary progressive aphasia
<sup>5</sup>lvPPA: logopenic variant primary progressive aphasia
<sup>6</sup>MoCA: Montreal Cognitive Assessment
<sup>7</sup>QAB: Quick Aphasia Battery<sup>30</sup>

#### 2.3.2 Methods

All PwPPA and care partners participated in the PACT protocol. Experimental sessions were conducted in person and audio- and video-recorded using a Panasonic Full HD Video Camcorder mounted on a tripod. Participants were directed in a structured conversation and responded to three standardized prompts:

1. <u>Case History:</u> Responses in conversation elicited by case history (e.g., “Can you tell me about the changes in communication you are experiencing?”).
2. <u>Light Conversation:</u> Participation in general or more personal conversation with clinician/conversation partner (e.g., “Can you tell you me about the work you do or a recent trip you have been on?”)
3. <u>Common Communication Tasks:</u> Explanations or demonstrations of common communication or conversational scenarios relevant to their daily lives (e.g., “Can you show me how you would make a phone call or write a short message to a loved one?”).

Care partners participated in the structured conversations, when present. Seven out of nine participants completed standardized cognitive-linguistic assessment during the same study visit through the UW ADRC or in the context of this experiment. Two participants declined and reported feeling discomfort and overburdened by the standardized measures. All participants were provided with the visual *Aphasia Impact Questionnaire – 21* (AIQ-21^29^) rating scale and asked to point to the image that corresponded to how they felt following standard cognitive-linguistic assessment and the PACT protocol, respectively. When possible, the *Quick Aphasia Battery* (QAB^30^) was used to screen for specific speech, language, and communication skills. Further open-ended participant feedback was solicited to contextualize participant ratings. Data collection occurred between June 26^th^, 2024, and November 10^th^, 2025.

#### 2.3.3 Results

Eight out of nine PwPPA rated the tool as superior to traditional neuropsychological testing. One participant with PwPPA reported greater fatigue during the PACT protocol than during standard pencil-and-paper testing due to the open-ended nature of the conversation. All six partners of PwPPA rated the tool as superior to traditional neuropsychological testing (see **Table 3** for PwPPA and care partner feedback).

**Table 3.** Reflections from PwPPA and care partners on the PACT protocol and traditional standardized assessment procedures. Participants were provided a 5-point rating scale (1= awful, 3 = fine, 5 = excellent) to evaluate testing procedures.

| **Participant** |  | **PACT** | **Traditional Assessment** |
| --- | --- | --- | --- |
| 1 | *PwPPA* | “Two thumbs up.” | “If it’s what you need.” |
|  | *Care Partner* | “[I give it a] 4 because the camera makes you self-conscious but paper testing is so tough... so this is better overall.” |  |
| 2 | *PwPPA* | “I am right in between [4 and 5]. I like talking. Communication is difficult. But I am here and want to connect.” | “Um. The testing reminds me that I’m degenerating. Impossible. I make a phone call as I told [coordinator] and they think I’m stupid or drunk. So, I feel this [pointing to 1.0] often.” |
| 3 | *PwPPA* | “So fast.” | [Hand wave] |
|  | *Care Partner* | “It didn’t even feel like testing. We have those conversations all the time anyway.” | “She feels defeated afterwards and it takes a while for her energy to go up again.” |
| 4 | *PwPPA* | “It takes...the longer I talk, the harder it gets. And of course open-ended questions like this one makes it harder. But yeah, I’d say this is typical. It feels like me.” | “Cognitive tests...that naming like naming animals or F words or things like that I can only get about 7 and 30 seconds and then just. Yeah. Even then it’s, that’s the naming part is the hardest part. Words I don’t use they want me to say.” |
|  | *Care Partner* | “That was so much easier and took much less time.” | “It was important for our diagnosis. But I don’t really see us needing that information again in the future.” |
| 5 | *PwPPA* | “I loved it. A conversation is what I needed.” | “No, thank you.” |
|  | *Care Partner* | “[The conversation] spoke to his humanity. He’s still there, and I’m not sure if drawing lines or naming hard-to-see pictures captures that.” | “It’s painful to watch him struggle.” |
| 6 | *PwPPA* | “Feels hard to do. It- because you’re asking me questions that uh... uh. That uh, that I used to have answers for, but don’t anymore. I used to have a facility with language. But I don’t anymore. And, and- um they ask questions. In my mind I could uh, I could reply. I have trouble.” | “Questions are closed and I can be short, so it feels easier. But I wouldn’t love to- to do it again.” |
|  | *Care Partner* | “Very natural and... efficient.” |  |
| **7** | *PwPPA* | “That felt really good. We had a really nice conversation. I give it a 4 because it won’t ever feel like a 5.” |  |
| **8** | *PwPPA* | “Well, it's it's...it doesn't take that much time and so much less time. And I don't know that writing things down, um. Writing things down, tests, I have all the time in the world to find that word. Talking to somebody like you in this I- I don't have the time. So it I either got I, I, I either grabbed the right word I wanted or I don't. And then I and I, I don't, I mean I just. I mean, I- I know that this is my life, so I just don't want to...sweat the small stuff. Waste my time. Hmm, so. Yeah, yeah, yeah, so this is easier in a way. And- and, but when I have to...communicate with somebody who's hard for me to communicate with...” | “I would say probably 3 like in the middle. It's. It certainly wasn't. Terrible. And it wasn't great. Umm, yeah, I mean it. I, I, I'm aware that I'm not going to get better. It helped me understand that. I'm, you know. Let's say slowly getting more challenged. Umm, so it was definitely. Challenging. Yeah, it, it just makes it more real. Yeah. And. Yeah. And and I'm somebody that is, I mean, I, I've been for sure living one day at a time and maximizing, you know that. And so I have. Very good ways that I'm never. Outboard. Mm-hmm. And if some amazing thing happens, well, but I don't think it's gonna happen in my time and I have no. Expectation that it will. Have some amazing reverse. I- I-, I mean, they were very kind the the people that I was working with this most recent time. It was, it was, I wouldn't go so far as to say shocking, but but it was a bit of a a wake up. Yeah, yeah. Imagine. It's why I'm. Being a little more. I mean. Getting my yeah my marbles... soon, whatever. I mean this is when I wish I could find what I want to say exactly, but- but yes it's. Yeah I'll just do my best.” |
| **9** | *PwPPA* | “Talking and finding the words, where I have time, and no pressure, makes this... I mean I feel great!” | “I try to say it, and then I think, [groan], this doesn’t feel right! I missed it already. And then it’s over.” |
|  | *Care Partner* | “This feels just like a natural conversation and it really speaks to his | “He gets performance anxiety. And so you |
|  |  | strengths that I see everyday.” | see him barely processing the information they give him and he can’t do it because he’s thinking about the fact that he might fail.” |

PwPPA on average rated their PACT protocol experience as 4.3 out of 5.0 (5 = excellent; SD: 1.4, range: 2.0-5.0, mode: 5.0) and that with the traditional neuropsychological assessment as 2.0 out of 5.0 (SD: 1.1, range 1.0-4.0, mode: 1.0). Care partners rated the PACT protocol experience as 4.8 out of 5.0 on average (SD: .44, range: 4.0-5.0, mode: 5.0) and neuropsychological assessment at 1.0 out of 5.0 (SD: 1.3, range: 1.0-4.0, mode: 1.0).

### 2.4 Experiment 4: Piloting the PACT with SLPs

#### 2.4.1 Participants

To explore acceptability and clinical implementability, 10 SLPs trialed the PACT to rate communicative strengths in PwPPA. Participants were recruited by word of mouth through the members of the International SLT/P PPA Network^26^ and National Aphasia Association.^31^ Inclusion criteria included licensure and experience in speech-language pathology, lack of participation in Experiments 1-3, and fluency in English. Participants reported an average of 10 years of clinical practice (SD: 3.6). Except for one, the clinicians practiced within the United States and cited English as their primary language of clinical practice. The other clinician practiced in Lebanon and identified Arabic as their primary language of clinical practice. For 9 participants, the target population was adult-onset neurogenic communication disorders. One participant described themselves as a generalist who worked across the lifespan. Eight participants identified subspecialty expertise in PPA and other neurodegenerative conditions. Participants were compensated with a $100 online gift card for their time.

#### 2.4.2 Methods

Participants met via Zoom with the first author (JG). Following consent, participants received an introduction to the purpose of the PACT and received a digital copy to review. After participants confirmed understanding of the study structure, participants watched a video recording of PwPPA participating in the PACT protocol. Participants were then asked to complete the PACT form based on this recording while verbalizing their thought process in doing so, following the ‘Think Aloud’^32,33^ procedure. These verbalizations included weighing ratings for each scale item, understanding (or lack thereof) of the wording, and overall thought processes as they navigated the PACT form while evaluating the PwPPA video. Each session was recorded to capture audio responses. Participants were then explicitly asked to describe their experience filling out the PACT form, identify its utility for clinical practice, indicate whether they would use it, and develop a tentative therapeutic plan for the videoed PwPPA based on the PACT protocol outcomes. Once completed, participants watched a second video of a different PwPPA completing the PACT protocol and given the option to complete the PACT ratings while watching, following the recording, or as before. The participants completed PACT ratings in a Microsoft Word document. Authors JG and JC evaluated the ‘Think Aloud’ solicited feedback as in Experiment 2. Data were collected between June 26^th^, 2025, and August 14^th^, 2025.

##### 2.4.2.1 PACT Stimuli

Four video recordings generated in Experiment 3 were used in this preliminary implementation phase. The recordings represented a person with svPPA (3min 0s), nfvPPA (6min 31s), lvPPA (7min 44s), and a mixed presentation (6min 55s). The svPPA and mixed presentation recordings included a care partner. Each recording had five raters. Recordings were counterbalanced to control for order effects.

#### 2.4.3 Results

Qualitative analysis of SLP feedback revealed four predominant areas of development. SLPs identified a need to digitize the PACT to enhance ease of usability, create domain or summary scores per PACT subscale, pull item-level descriptions into documentation, and consider that a person’s baseline performance may not be reflected in the PACT scoring system. No changes to item-level content of the four PACT scales were recommended. See **Supplementary Table 3A** for comprehensive feedback and coding.

### 2.5 Experiment 5: Piloting the Digital PACT with SLPs

Feedback from the initial validation phase was incorporated to create the final version of the PACT. The first author proposed revisions and presented these to the expert panel defined in Experiment One. These revisions addressed the format of the outcome measure, the instructions, and the frequency and placement of open-ended clinician observations, self-report, and partner-reported strengths. These changes were intended to streamline instructions and eliminate open-ended items in each subscale, replacing them with single instances of clinician observations, self-report, and self-reported strengths. The PACT was coded by author JG into a Shiny app,^34^ a user interface coded in R statistical language, to ease public accessibility and usability of the tool: https://jgallee.shinyapps.io/pact/.

#### 2.5.1 Participants

Three additional participants were recruited through the International SLT/P Network. Exclusion criteria included participation in any element of the PACT process described in Experiments 1-4. As in Experiment 4, inclusion criteria included active clinical practice and experience with adults with acquired neurogenic communication disorders. The participants were all female, native English speakers practicing in the USA and UK. The average clinical practice experience was 19.7 years (SD: 15.5).

#### 2.5.2 Methods

The three participants completed the same procedure as described in Experiment Four with the updated PACT format. Participants were provided with a link to the Shiny web application to complete the PACT protocol after consent. Participant feedback on the clinical usability of the PACT was qualitatively analyzed by authors JG and JC. Data collection occurred between September 15^th^, 2025, and October 22^nd^, 2025.

#### 2.5.3 Results

##### 2.5.3.1 Experiment 5: Digital Efficiency

Participants took an average of 20 minutes (SD: 3.6) to complete the scale the first time while verbalizing their thought processes. The average time to completion at the second attempt was 12.5 minutes (SD: 13.3).

##### 2.5.3.2 Experiments 4 and 5: Agreement on Measurability and Strength

All 13 SLPs from Experiments 4 and 5 affirmed that the PACT protocol was sufficient to evaluate communication and provide preliminary intervention recommendations using a single conversational sample. There was 91% agreement on the overall measurability of the PACT items and 85% agreement on the evaluation of strength for the four recordings across all raters (n = 13; see **Table 5 and Supplementary Table 4**). There was 92% agreement on measurability for recordings that included a care partner and 91% on recordings without. *Speech and Voice* had the highest overall agreement on measurability (100%), followed by *Discourse* (94%), *Social-Pragmatics* (92%), and *Language* (89%). Strength agreement was highest for *Language* (85%), followed by *Speech and Voice* (85%), *Social-Pragmatics* (84%), and *Discourse* (73%). Apart from *Social-Pragmatics* (Mixed: 73%), agreement on measurability across domains and diagnoses was above 85%. Percent agreements of strength evaluation were upwards of 80% for domains by diagnosis, with notable exceptions for *Speech and Voice* (nfvPPA: 59%), *Social-Pragmatics* (svPPA: 62%, Mixed: 66%), and *Discourse* (svPPA: 47%, Mixed: 68%). See **Supplementary Table 4** for all measures of agreement.

**Table 4.** Participant demographics for Experiment Two: Phases Two and Three (piloting the completion of the PACT).

| **Participant** | **Gender Identity** | **Years of Clinical Practice** | **Primary Patient Population** | **Country of Practice** | **Language of Practice** |
| --- | --- | --- | --- | --- | --- |
| 1 | Female | 10 | Acquired adult neurogenic communication disorders; PPA; neurodegenerative conditions | USA | English |
| 2 | Female | 5 | Across the lifespan | USA | English |
| 3 | Female | 10 | Acquired adult neurogenic communication disorders; PPA; neurodegenerative conditions | USA | English |
| 4 | Female | 19 | Acquired adult neurogenic communication disorders; PPA; neurodegenerative conditions | USA | English |
| 5 | Female | 10 | Acquired adult neurogenic communication disorders; PPA; neurodegenerative conditions | USA | English |
| 6 | Female | 7 | PPA; neurodegenerative conditions | USA | English |
| 7 | Female | 11 | Acquired adult neurogenic communication disorders | USA | English |
| 8 | Female | 12 | Acquired adult neurogenic communication disorders | Lebanon | Arabic; English |
| 9 | Female | 10 | Acquired adult neurogenic communication disorders; PPA; neurodegenerative conditions | USA | English |
| 10 | Female | 10 | Acquired adult neurogenic communication disorders; PPA; neurodegenerative conditions | USA | English |
| 11 | Female | 15 | Acquired adult neurogenic communication disorders; PPA; neurodegenerative conditions | US | English |
| 12 | Female | 7 | Acquired adult neurogenic communication disorders; PPA; neurodegenerative conditions | US | English |
| 13 | Female | 37 | PPA | UK | English |

**Table 5.** Ratings of PACT item importance and pilot inter-rater agreement. Delphi respondents evaluated items on a scale from 1 to 5 (1 = not at all important, 3 = somewhat important, 5 = very important). Ten speech-language pathologists scored PwPPA participating in the PACT protocol, prior to the launch of the web-based application. Each of the ten raters completed the Microsoft Word form of the PACT form on two videos, resulting in five ratings per recording. Raters followed the PACT strengths-based scale (4 = consistent strength, 3 = intermittent strength, 2 = limited strength, 1 = not a strength, 0 = not evaluated).

| DOMAIN | ITEM |  | DELPHI OUTCOMES |  |  | INTER-RATER AGREEMENT [weighted]* |  |  |  |
| --- | --- | --- | --- | --- | --- | --- | --- | --- | --- |
|  | # | Stimulus | Mean | SD | Mode | svPPA | nfvPPA | lvPPA | Mixed |
| General | 1 | Clinician-observed personal strengths (*address use of alternative modalities for input/output, e.g., gestures, writing, key wording, photos, and applications*) | 4.7 | .70 | 5.0 |  |  |  |  |
|  | 2 | Self-reported strengths | 4.8 | .53 | 5.0 |  |  |  |  |
|  | 3 | Partner-reported strengths | 4.7 | .62 | 5.0 |  |  |  |  |
| Speech and Voice | 1 | Voice/Resonance (*breathiness/breath support, loudness*) | 4.0 | .91 | 5.0 | .8<br>[1] | .6<br>[.6] | 1<br>[1] | .8<br>[.8] |
|  | 2 | Articulatory precision (*absence of speech sound distortions*) | 4.2 | .93 | 5.0 | .8<br>[1] | .6<br>[.6] | .4<br>[.8] | 1<br>[1] |
|  | 3 | Word form accuracy (*absence of speech sound errors whether due to phonological or motor speech concerns*) | 4.5 | .88 | 5.0 | 1<br>[1] | .6<br>[.6] | .6<br>[.8] | 1<br>[1] |
|  | 4 | Word production and flow (*absence of false starts, perseverations, and significant pausing whether due to phonological or motor speech concerns*) | 4.3 | .87 | 5.0 | .6<br>[1] | .6<br>[.6] | .8<br>[.8] | .8<br>[1] |
|  | 5 | Ease of production (*absence of perceived motoric effort*) | 4.2 | 1.1 | 5.0 | .8<br>[.8] | .8<br>[.8] | .6<br>[1] | .4<br>[.8] |
|  | 6 | Use of prosody and intonation to enhance communicative intent (*pitch and timing cues and variation to indicate emotion/emphatic stress, (dis)agreement, or grammatical relations*) | 4.0 | 1.0 | 5.0 | .8<br>[1] | .6<br>[.6] | .8<br>[1] | .4<br>[.6] |
|  | 7 | Rate of speech (*non-obstructive: not too slow or too fast*) | 3.9 | 1.2 | 5.0 | .6<br>[1] | .8<br>[.8] | .6<br>[1] | .4<br>[.8] |
|  | 8 | Intelligibility (*overall ease with which the communicator is understood when communicating via spoken language*) | 4.5 | .88 | 5.0 | 1<br>[1] | .4<br>[.8] | .6<br>[1] | .8<br>[1] |
| Language | 1 | Specificity of word retrieval (*the opposite of non-specific language, e.g., “thing” or “stuff”*) | 4.5 | .59 | 5.0 | .6<br>[.6] | .8<br>[1] | .6<br>[.6] | .8<br>[1] |
|  | 2 | Semantic accuracy (*ability to convey word meaning accurately or appropriateness of word choice as it relates to meaning, even if non-specific*) | 4.7 | .56 | 5.0 | .6<br>[.8] | .8<br>[1] | .6<br>[1] | 1<br>[1] |
|  | 3 | Syntactic complexity (*relative complexity of phrases/sentences in terms of type and length*) | 4.4 | .81 | 5.0 | .4<br>[0.6] | .6<br>[0.8] | .4<br>[0.8] | .6<br>[1] |
|  | 4 | Informativeness/topic completeness (*completeness and explanatory power of message communicated*) | 4.5 | .51 | 5.0 | .6<br>[0.8] | .8<br>[1] | .6<br>[1] | .6<br>[1] |
|  | 5 | Circumlocution (*communicator's success in speaking around a topic or find alternative methods to describe them*) | 4.4 | .66 | 5.0 | .6<br>[.6] | .6<br>[1] | .4<br>[.6] | .8<br>[.8] |
|  | 6 | Comprehension of spoken single words (*concepts or actions*) | 4.7 | .55 | 5.0 | .8<br>[.8] | .6<br>[.6] | .6<br>[.6] | .6<br>[.6] |
|  | 7 | Comprehension of spoken phrase-level output (*statements, commands, or questions*) | 4.6 | .65 | 5.0 | .6<br>[.8] | 1<br>[1] | .6<br>[1] | 1<br>[1] |
|  | 8 | Use of text-based AAC<sup>1</sup> to augment spoken output and input (*e.g., writing, reading, text-to-speech generators in a single conversational turn*) | 4.3 | .76 | 5.0 | 1<br>[1] | .6<br>[1] | .6<br>[.6] | .4<br>[.4] |
|  | 9 | Use of pictures or iconographic AAC to augment spoken output and input (*in a single conversational turn*) | 4.2 | .88 | 5.0 | 1<br>[1] | .6<br>[.8] | .8<br>[.8] | 1<br>[1] |
| Social-Pragmatics | 1 | Participation in communicative context (*spoken or other engagement and initiation in communication, including but not limited to conversational turn-taking, and initiating relevant topics or ideas*) | 4.6 | .65 | 5.0 | .4<br>[.8] | 1<br>[1] | .8<br>[1] | .8<br>[1] |
|  | 2 | Initiation of spoken or compensatory communication repair strategies (*independent implementation of strategies to smooth over communication breakdowns*) | 4.5 | .59 | 5.0 | .4<br>[.4] | .4<br>[.8] | .4<br>[.6] | .4<br>[.4] |
|  | 3 | Use of spoken or compensatory communication repair strategies when provided support (*supported implementation of strategies to smooth over communication breakdowns*) | 4.4 | .58 | 4.0 | .6<br>[.6] | .6<br>[.6] | .8<br>[.8] | .6<br>[.6] |
|  | 4 | Social appropriateness of participation (*including but not limited to mirroring body language, maintaining expected comportment and engagement with clinician, and purposeful adherence to expectations and sharing of content in communication*) | 4.4 | .82 | 5.0 | .6<br>[.6] | 1<br>[1] | .8<br>[1] | .8<br>[1] |
|  | 5 | Perceived empathy or sensitivity to communication | 4.2 | .92 | 5.0 | .4 | 1 | .6 | .6 |
|  |  | partner (*recognition and responsiveness to clinician/conversation partner and in terms of topic content*) |  |  |  | [.6] | [1] | [.8] | [.6] |
|  | 6 | Use of body language to emphasize, refer, or explain objects, events, and actions (*use of gestures, enactments, or visualizations to communicate intended meaning*) | 4.3 | .87 | 5.0 | .4<br>[.6] | 1<br>[1] | .6<br>[.8] | .6<br>[.6] |
|  | 7 | Use of facial expression to enhance communication (*e.g., to communicate internal states or feelings about content of a topic or situation*) | 4.3 | .86 | 5.0 | .6<br>[.6] | 1<br>[1] | .8<br>[1] | .6<br>[.8] |
|  | 8 | Strategic competence in the use of communication supports/AAC to flexibly switch between communication modalities to communicate an intended message and maintain dialogue with clinician/conversation partner | 4.5 | .66 | 5.0 | .8<br>[.8] | .6<br>[.8] | .8<br>[.8] | .6<br>[.6] |
| Discourse | 1 | Establishing of topic (*does the communicator clearly introduce their target topic?*) | 4.3 | .81 | 5.0 | .4<br>[.6] | 1<br>[1] | .4<br>[.8] | .8<br>[.8] |
|  | 2 | Confirmation of listener comprehension (*e.g., does the communicator identify moments of misunderstanding when appropriate?*) | 4.2 | .92 | 5.0 | .4<br>[.6] | .4<br>[.8] | .6<br>[.6] | .8<br>[.8] |
|  | 3 | Topic relevance (*as based on context and/or prompt*) | 4.3 | .76 | 5.0 | .4<br>[.6] | .8<br>[1] | .6<br>[.6] | .8<br>[.8] |
|  | 4 | Inclusion of story grammar elements (*characters/agents, setting, actions, resolutions, and more*) | 4.0 | .86 | 4.0 | .8<br>[.8] | .8<br>[1] | .6<br>[1] | .6<br>[.6] |
|  | 5 | Cohesion and coherence (*logical flow of utterances and ideas*) | 4.5 | .83 | 5.0 | .6<br>[.6] | .8<br>[1] | .6<br>[.8] | .4<br>[.8] |
|  | 6 | Efficiency (*how many attempts, how much time, or overall quantity of output is required to communicate the intended message?*) | 4.3 | .76 | 5.0 | .4<br>[.6] | .6<br>[1] | .6<br>[.6] | .6<br>[.6] |
| **Averages (SD)** |  |  | **4.4**<br>(.21) |  |  | **.6**<br>[.8] | **.7**<br>[.9] | **.6**<br>[.8] | **.7**<br>[.8] |

\*Recoded to collapse strength ratings as strengths (consistent strength and intermittent strength), not strengths (limited strength and not a strength), or not evaluated.
<sup>1</sup>AAC: *Augmentative and Alternative Communication*

##### 2.5.3.3 Experiments 4 and 5: Reflections on Clinical Feasibility, a Holistic and Strengths-Based Approach, and Supporting Use for Practice

Qualitative feedback from Phases Two and Three of Experiment Two revealed three major themes: clinical feasibility and utility, a focus on identifying and leveraging strengths, and barriers/enablers that support use in practice. Examples are provided below (see **Supplementary Table 3B** for more).

#### 2.5.3.3.1 Clinical Feasibility and Utility

Participants expressed strong support for the clinical utility of the PACT, highlighting that it was feasible to implement.

“I think from like a teaching and a clinical use perspective, it is so deeply needed.”

“I’m really confident I could absolutely do this after doing an evaluation with someone or having a couple of conversations with them.”

“I’m really interested and excited actually to use it! I think it’s much more intuitive… going through it.”

“If I’m in university clinic… I am like making each one of my students, you know, fill this kind of thing out.”

“It captures kind of a wide variety of communication so that you can really showcase like what other areas where they are really strong and when they’re doing a really fantastic job … as a clinician, I would absolutely like this.”

#### 2.5.3.3.2 A Focus on Finding and Leveraging Strengths

Participants stated that the PACT provides rich, holistic information and reframes assessment as real-world support by offering an alternative to standardized assessment.

“It’s helping me think about things in a little bit of a different light instead of deficit-based, which is usually what we’re thinking about…. What can this person do? What’s a strength and what’s not a strength?”

“I especially like…having your thinking set up or not you know what are the deficits…what can we actually do about it? I think making that gap is…where that like clinical kind of creativity or you know, the magic in therapy happens.”

“I definitely feel like it was comprehensive but fairly quick and got the wheels spinning in a way that I don’t think I would necessarily would have thought otherwise.”

“It could be widely applied to neurologists or other primary care providers even. Because obviously the key here is to have early detection and to refer them to specialist centers as soon as possible for intervention.”

#### 2.5.3.3.3 Barriers and Facilitators

Participants identified barriers and facilitators of supporting the use of the PACT in practice. These points touched upon the necessity of a recording, the logistics of use, time, flexibility, setting, and training.

“Depending on my setting, the ability to [record] is limited.”

“There would be at least some of your users that … wouldn’t have access to the app.” “[A] quick scale …that is not too labor intensive.”

“I could very much see myself … pulling it up and referencing and like using it as a benchmark tool.”

## 4 DISCUSSION

In this project, we set forth to develop an inventory of communication, structured as a set of rating scales that allow clinicians to use a common framework to synthesize speech, language, and communication function. We collaborated with partners in research to develop an instrument sensitive to PPA that generates a comprehensive communication profile from structured conversational prompts that engage the client, care partner, and clinician. The PACT captures listener perceptions of a person’s communication and contextualizes these findings with qualitative insights from PwPPA, care partners, and the clinician. The PACT was born out of the desire to reframe communication ability in PwPPA, minimize assessment burden, facilitate referrals to intervention, and shape intervention. Moreover, the PACT leverages the conversations that PwPPA and care partners are likely to have with most clinical providers or researchers while minimizing the discomfort of being tested. Pilot feedback is highly promising and indicates that the PACT meets these aims for PwPPA with mild to moderate severity of symptoms.

Communication is central to human connection. Its richness extends beyond linguistic accuracy. Beyond words, a range of behaviors can sustain communication success even in the presence of language decline. Recognizing these strengths is critical for promoting participation and well-being. The PACT enables clinicians and researchers to document and monitor symptom-led communication strengths and treatment responsiveness through structured conversation. This approach centers on what a person *can* do and supports clinical decision-making for individualized, person-centered intervention.

### 4.1. Future directions

Future directions include instrument validation and the establishment of normative parameters for PACT outcomes. We will also further evaluate the PACT’s reliability and utility in shaping intervention goals through clinical provider feedback and modify items as needed to improve the scales. Finally, we will meet the calls for globally responsive tools^35–37^ and collaborate with clinical researchers worldwide to develop cultural-linguistic adaptations.

## Data Availability

All data produced in the present study are available upon reasonable request to the authors.

## 6 ACKNOWLEDGEMENTS

We are deeply grateful to the people living with PPA and their families who generously dedicated their time, experiences, and energy to this work, thereby providing the foundation for advancing our understanding of PPA and improving clinical care.

## 9 CONSENT STATEMENT

All human subjects involved in the present study provided written informed consent.

