## Supplementary Table 1 for "The Progressive Aphasia Communication Toolkit (PACT): A Strengths-Based Approach to Multidomain Evaluation for Intervention"

**Supplementary Table 1.** The members of the expert panel responsible for the original draft of the instrument in Experiment 1, prior to the Delphi consensus process in Experiment 2.

| **Participant** | **Research Department** | **Clinical Profession*** | **Area(s) of Expertise** | **Population of Expertise** | **Work Setting** | **Location** |
| --- | --- | --- | --- | --- | --- | --- |
| Jeanne Gallée, PhD, CCC-SLP | General Internal Medicine | Speech-Language Pathologist | Assessment; life participation approaches | ADRD^1^  PPA^2^ | University | Seattle, Washington, USA |
| Jade Cartwright, PhD | Health Sciences | Speech Pathologist | Psychosocial intervention; life participation approaches | ADRD^1^  PPA^2^ | University | Hobart, Tasmania, Australia |
| Maya L. Henry, PhD, CCC-SLP | Speech, Language, and Hearing Science | Speech-Language Pathologist | Rehabilitation | PPA^2^ | University | Austin, Texas, USA |
| Aimee R Mooney, MS, CCC-SLP | Neurology | Speech-Language Pathologist | Geriatrics; rehabilitation | PPA^2^ | University Hospital | Portland, Oregon, USA |
| Brielle C Stark, PhD | Speech, Language, and Hearing Sciences | Researcher | Connected speech and discourse | Aphasia | University | Bloomington, Indiana, USA |
| Anna Volkmer, PhD | Language and Cognition | Speech Therapist | Communication partner training; rehabilitation | PPA^2^ | University | London, UK |
| Rob J Fredericksen, PhD | Allergy & Infectious Diseases | Researcher | Patient reported outcomes; behavioral health; qualitative analysis | HIV | University | Seattle, Washington, USA |
| Kimiko Domoto-Reilly, MD, MMSc | Neurology | Neurologist | Neurological assessment and care | ADRD  PPA  FTD^3^ | University; Memory Clinic | Seattle, Washington, USA |
| Paul K. Crane, MD, MPH | General Internal Medicine | General Internist | Psychometrics; patient reported outcomes | ADRD^1^ | University; Hospital | Seattle, Washington, USA |

*as described in the country of practice

^1^ADRD = Alzheimer’s disease and related disorders

^2^ PPA = primary progressive aphasia

^3^ FTD = Frontotemporal degeneration spectrum diseases
