## Supplementary Tables 2A and 2B for "The Progressive Aphasia Communication Toolkit (PACT): A Strengths-Based Approach to Multidomain Evaluation for Intervention"

**Supplementary Table 2A. Original instrument prototype generated in Experiment 1.** The prototype was named the Functional Communication Checklist (FCC). The authors were Jeanne Gallée, PhD; Jade Cartwright, PhD; Maya L Henry, PhD; Aimee Mooney, MS; Brielle C Stark, PhD; Anna Volkmer, PhD; Connie Nakano, MS; Rob J Fredericksen, PhD; Kimiko Domoto-Reilly, MD, MMSc; Paul K Crane, MD, MPH.

| **DOMAIN** | **ITEM** |
| --- | --- |
| Speech | Voice/Resonance  (breathiness/breath support, loudness) |
|  | Articulatory precision (clarity of target speech sounds; speech distortions) |
|  | Word form accuracy  (do the words have speech sound errors?) |
|  | Word production  (absence of false starts and perseverations) |
|  | Flow (absence of significant pausing) |
|  | Prosody (use of pitch, phrase boundaries, and lexical stress) |
|  | Rate of speech |
|  | Clinician-observed strengths |
|  | Self-reported strengths |
| Language | Specificity of word retrieval (does the speaker use underspecified/vague/empty language or specific/meaningful to refer to target words or concepts) |
|  | Semantic accuracy (accuracy or appropriateness of word choice as it relates to meaning) |
|  | Syntactic complexity (relative complexity of phrases in terms of type and length) |
|  | Informativeness/topic completeness (does the speaker’s point come across?) |
|  | Circumlocution (does the speaker successfully speak around a topic or find alternative methods to describe them?) |
|  | Comprehension of single words (concepts or actions) |
|  | Comprehension of phrase-level output (statements, commands, or questions) |
|  | Clinician-observed personal strengths |
|  | Self-reported strengths |
| Discourse | Establishing of topic (does the speaker clearly introduce their target topic?) |
|  | Topic relevance (as based on context and/or prompt) |
|  | Inclusion of story grammar elements (characters/agents, setting, actions, resolutions, and more) |
|  | Cohesion and coherence (logical flow of utterances and ideas) |
|  | Efficiency (how long does it take the speaker to communicate an intended idea using any modality?) |
|  | Functional success in interaction (success and efficacy in communicating an intended message based on typical daily activities; multiple should be used for scoring): Examples   1. Could you show me how you’d make a phone call to a loved one? 2. How would you order your typical meal at your favorite restaurant? 3. Please show me how you’d respond to an email or text message from a friend. 4. Please show me how you’d look up your doctor’s address. 5. Can you explain what’s troubling you about your language to me? |
|  | Clinician-observed personal strengths |
|  | Self-reported strengths |
| Cognition | Initiation (purposeful and independent initiation of communicative participation) |
|  | Inhibition (purposeful, voluntary restraint and adherence to expectations and sharing of content in communication) |
|  | Perception (identification and processing of stimuli in immediate environment) |
|  | Selective attention (attention to conversation partner and tasks) |
|  | Sustained attention (maintenance of attention to conversation partner and tasks in this context) |
|  | Short-term memory (maintenance and use of information relevant to current context) |
|  | Long-term memory (maintenance, retrieval, and use of information prior to current context) |
|  | Clinician-observed personal strengths |
|  | Self-reported strengths |
| Social-Pragmatics | Participation in communicative context (engagement and initiation in communication, including but not limited to responding to the clinician, conversational turn-taking, and initiating topics or ideas) |
|  | Initiation of communication repair strategies (independent implementation of strategies to smooth over communication breakdowns) |
|  | Use of communication repair strategies when provided support (supported implementation of strategies to smooth over communication breakdowns) |
|  | Social appropriateness of communication or participation (including but not limited to mirroring body language, maintaining expected comportment and engagement with clinician)) |
|  | Empathy or sensitivity to communication partner (recognition and responsiveness to clinician as a human and in terms of topic content) |
|  | Use of body to explain or refer to objects, events, and actions (use of gestures, enactments, or visualizations to communicate intended meaning) |
|  | Use of facial expression to enhance communication (use of facial expressions to communicate emotional state or feelings about content of a topic or situation) |
|  | Use of prosody and intonation to enhance communicative intent (pitch and timing cues to indicate emotion, (dis)agreement, or grammatical content) |
|  | Use of communication support to enhance communication (support is defined as AAC^1^, writing, drawing, pointing to objects, low and high tech (including but not limited to pictures, word books, and smart phones: also evaluate the strategic competence in flexibly switching between communication modalities to communicate an intended message) |
|  | Clinician-observed personal strengths |
|  | Self-reported strengths |

*^1^AAC: Augmentative and Alternative Communication*

**Supplementary Table 2B. Summarized feedback on the Functional Communication Checklist (FCC) from each of the Delphi Rounds in Experiment 2.** This information was presented to the electronic Delphi survey respondents in Rounds 2-4, in addition to the item-level ratings (see **Table 1** in main body of text). The feedback summary column reflects authors JG and RJF’s coding of the broad themes of changes requested by survey respondents following each round.

| **Round** | **Remain As Is** | **Feedback Summary** | **Approach** |
| --- | --- | --- | --- |
| **1** | **66%** | **34%** provided constructive open-ended feedback to enhance the proposed instrument. Broadly, this feedback could be broken down into the following themes: 1) clarify item-level definitions 2) clarify terms (e.g., functional communication and item-level prompts) 3) clarify *how* the FCC is to be used: its overarching purpose and which prompts should be used to fill out the FCC 4) incorporate care partner evaluation into the scoring 5) integrate communication modalities across the board into the FCC | **Instructions** Our first change was to include a complete set of instructions on how to implement the FCC. This also includes concrete definitions of what we mean by functional communication and the goals of the FCC. Furthermore, we have integrated and specified the importance of care partner participation and input in completing the FCC (when available).  **Rating Scale** We have also updated the rating scale to be more consistent with our strengths-based approach. Previously, we used a 3-point scale (1 = mild, 2 = moderate, 3 = severe) to rate interference with functional communication.  **Modalities** We previously neglected to systematically address the modalities we hope to assess and capture in using the FCC. We have also integrated and clarified which modalities we'd like the clinician to examine and evaluate when using the FCC.   **Items** In the open-ended feedback we received, survey respondents listed the following concerns: 1) lack of care partner input 2) lack of clarity of which modalities are evaluated 3) insufficient attention drawn to the use of AAC and other compensatory strategies 4) lack of clarity in certain item-level definitions  We have made distinct changes throughout the scale to address these concerns. These include, but are not limited to, more appropriate conversational prompts to complete the FCC (which is now no longer an item in the Discourse section but in the instructions), enhanced item-level definitions, collapsing of items that are too similar in nature, the addition of items addressing patient fatigue and perceived effort, as well as additional items to evaluate the use of specific types of alternative communication modalities). We hope to have sufficiently addressed your feedback while remaining true to the aims of the FCC. |
| **2** | **90%** | **10%** provided constructive open-ended feedback to enhance the proposed instrument. Broadly, this feedback could be broken down into the following objectives: 1) restructuring the instructions to enhance clarity and define augmentative and alternative communication (AAC) 2) reduce the overlap between items in the cognition and social-pragmatics sections 3) adding a text-box to capture the "greatest challenge" for a client | **Instructions** Our first change was to include an example scenario in the instructions that we included in Version 2.  This also includes a concrete definition of alternative and augmentative communication (AAC).   We also updated the instructions to be more clearly structured.   **Order** Based on feedback, we have reordered the survey sections of to reflect that ratings are based on conversational prompts and the order in which a clinician may evaluate a client's performance/input. The original order was speech, language, discourse, cognition, and social-pragmatics. The current order is discourse, social-pragmatics, language, speech, and cognition.    **Items** We reduced some of the items to avoid overlap between cognition and social-pragmatics, resulting in the items "initiation" and "inhibition" being removed from the cognition section. Furthermore, we updated the wording of some of the items to ensure their distinct purpose.   **Terminology** In our item descriptions, the concept of "verbalization" has been updated to reflect our true intention: spoken language.   As in the previous round, we hope to have sufficiently addressed your feedback while remaining true to the aims of the FCC. |
| **3** | **95%** | **5%** provided constructive open-ended feedback to enhance the proposed instrument. Broadly, this feedback could be broken down into the following objectives: 1) remove the section on cognition 2) refine the purpose and background to clarify motivation and usefulness for therapy | **Name** Upon great reflection, we have made the decision to move away from the "Functional Communication Checklist" to the "Progressive Aphasia Communication Toolkit", or **PACT**. Our tool was born out of the desire to promote functional communication in our clients. This scale does not measure functional communication, but rather is a comprehensive inventory of communication and its subdomains that can be used to create functional targets in intervention. As such, the tool is now renamed.  Why Toolkit? The instrument is a collection of scales. One of the "arcs" of this work is to add in handouts for care partners and people living with PPA (and related communication changes) to help support communication at home. Furthermore, it is a tool for assessment that should be used to help shape therapy and to think about our clients holistically. As such, the current working name is the PACT. This also allows us to extend this work to diagnoses outside of PPA. There is now a decision-tree for any clinician making choices about only filling in one, rather than all, of the scales.  **Instructions** The instructions have been further refined and shortened. The instructions also speak to the purpose of the PACT: to document outcomes that help support person-centered intervention. Here, the prompt box for the clinician to fill out during assessment has also been enhanced. The three standardized topics of conversation have been included in the text box and call for prompts on (1) case history/communication challenges, (2) light conversation (e.g., small talk), and (3) demonstration of a person-specific communication task. The instructions also speak to how ratings should be completed (e.g., following the video recording to allow the clinician to remain present in the interaction).  **Domains** The instrument now consists of *four*, rather than *five* scales. Cognition is now no longer specifically assessed. This is because the actual assessment consists of a conversation and no actual standardized assessment of cognition takes place. The instructions note that all results should be contextualized by outcomes of independently collected assessments of cognition.  **Items** We have further reduced items to avoid redundancy, as well as further clarified item descriptions. The scale originally named "Speech" has now been renamed "Speech and Voice" to better describe the item content.   **Scale** The rating scale has been updated to reflect our mission to create a strengths-based instrument. Now, the items are rated on a 5-point Likert scale from 4-0, where 4 = consistent strength, 3 = intermittent strength, 2 = limited strength, 1 = not a strength, and 0 = not evaluated.   As in the previous round, we hope to have sufficiently addressed your feedback while remaining true to the aims of the PACT. |
| **4** | **98%** | The sole theme identified was the possible digitizing of the instrument | **Changes halted due to response saturation and transition to piloting in Experiments 3 and 4.** |
