## Supplementary Table 3A and 3B for "The Progressive Aphasia Communication Toolkit (PACT): A Strengths-Based Approach to Multidomain Evaluation for Intervention"

**Supplementary Table 3A. Qualitative feedback from speech-language pathologists piloting the PACT assessment form based on two video recordings of PwPPA completing the PACT protocol.** Feedback was drawn from the ‘Think Aloud’ protocol. Speech-language pathologists were asked to discuss the clinical usability and feasibility of the PACT within their current assessment practices. Participant feedback led to the development of the digitized PACT to enhance usability, consolidate item-level descriptions into a table readout, create summary scores for each PACT subscale, and note that a person’s baseline performance may not be reflected in the PACT scoring system.

| **Participant** | **Enhancing Usability** | **Summarize Scores** | **Pull Item-Level Information into Documentation** | **Baseline Performance** |
| --- | --- | --- | --- | --- |
| 1 | “Yeah, I think I'm of two thoughts. One is depending on my setting, the ability to use recording is limited, yes. So if I'm really honoring what the scale is asking me to do, which is to watch a recording afterwards, that's a meeting. And probably more likely to use something live, but I do think especially if I'm being able to use umm, you know that quick scale or click a button or do something that is not too labor intensive than I would nice. Um, and I also think like even if you don't have the recording, it is the kind of thing that you could fill out relatively easy. After they’re when they're not live with you.”  “And I think what I had been using for sort of kind of almost the same thing, but not really in the same lens as the [Progressive Aphasia Severity Scale]. Right. Umm, which I like, but it's so hard to see and read because you're trying to like, you know, it's the same kind of, it's the same challenge, right? Of like if you're going to look at all the different domains and be specific about each of the characteristics, then it obviously has to be either multiple sheets or. Yeah, I don't know how you get it all on one page.”  “And again, I know that this is like in a paper format now and it's not necessarily meant to be, but I think that there would be at least some of your users that would be having, they wouldn't have access to the app, right? Again, because of like a hospital, you know, use or if you're like going into like, well, I'm going to take a step back and say you're probably not going to be doing this in a hospital like acute care setting. If you're an outpatient, like you're gonna have some different options, but I don't love when I have to wrestle through a ton of my papers and I tend to forget to write down, um. My own little personal notes or I'm like writing them very fast or very abbreviated and trying to get it into the computer screen quickly.” | **“**If you have like sort of a template of what would that look like, a section or like some sort of like scoring sheet. But if there's like a, a synopsis, you know, kind of like, now I have all this information and. I have a more structured um. Report, umm. And so that it can live it out into something that's, yeah, structured, easy to glance at.” | “Section would have something like continue to monitor these things for areas of strength and it might also, you know depending on if I’ve done other rating scales or how much I've gotten from that probably from that case interview right there probably would be a ton of information about like her personal strengths or partner strengths. So then I would put that in there too and potentially. From a goal around confidence or use of compensatory strategies based on the information we just gathered.”  “You have all this beautiful description that I'd want to be able to capture in, but I'm not necessarily going to retype it…I would probably pull snippets forward into like a formal reports, S.O.A.P note, use it more as a therapeutic tool and then also as a cross time benchmarks that I can show change.” | **“**How much [performance] is outward facing versus how much is just for me to look at and then make, you know, those kind of broader statements?” |
| 2 |  | **“**But yeah, I think for me, I tend to be a lot more condensed in my information and I'm going to be taking notes on this stand out things, not necessarily section by section.”  “Yeah, I think what I want is I want like almost like add up the scores. Then say like to say like, you know, I always want the numbers to help guide me towards like this. You know, you have it there. But like if it's a 3.5, like this is a relative strength or you know, whatever it is. So like a totality of, I mean, I could do that myself. That if there is a prompt in there to have like an average of the scores for each domain separately or overall yeah, each domain separately.” | “The one pager where you can put your cumulative thoughts. I think it could be a product of my like newness to this form, but for me probably just like one box where I could just take notes. Kind of what I did here, which was like I put all my notes in the top box and then I like would then put them in the appropriate places later on.” | “What's a consistent strength to not a strength at all? Like even examples of those would be useful once you start to get those norms. Because also, like when I was watching the video, I was thinking of the difference between the husband and wife. Like men and women speak differently oftentimes, right? So like, what's the what's the norm for a man versus a woman, right? His wife had plenty of pauses, but she included lots of details. But if you're a sparse communicator at baseline, then I'm dinging you for things that. Or just your baseline.” |
| 3 | “Yeah, probably 1 box, the top for notes and then the the clinician self report, partner report strength sections. Maybe it would be like if, if that information is really important to have for each of them, maybe then like additional problems at the very end, like OK, can I just ask you like what are some strengths that you see in yourself or? Your speech and voice maybe that's too explicit but then seeing if they have a response.” |  |  |  |
| 7 |  | “I do think that's a valuable addition to have. I, I do think that once I have those written down with like their respective places with the, the specific sections of like the, the domains. I think that will possibly like have a a more holistic idea or like it will give a better representation of the participant yeah for sure I didn't thank you for asking that question because I had forgotten that those are part of it I was just like so focused on the [individual] scores.” |  |  |

**Supplementary Table 3B. Qualitative feedback from speech-language pathologists piloting the PACT from the ‘Think Aloud’ protocol.** Authors coded participant feedback as *Clinical Usability and Feasibility, a Focus on Finding and Leveraging Strengths,* and *Barriers and Facilitators*.

| **Participant** | **Clinical Usability and Feasibility** | **A Focus on Finding and Leveraging Strengths** | **Barriers and Facilitators** |
| --- | --- | --- | --- |
| 1 | “If I'm in university clinic, ohh, heck yeah. So I am like making each one of my students, you know, fill those kind of thing out. And I also like a thinker especially I think with progressive illness do tend to be more interested in like trying to like put symptoms or put areas of relative strength into some sort of grid or some sort of paper format and then chart it and keep it across time. So I could very much see myself having like a paper folder or a file storage, you know, on my computer. And pulling it up and referencing and like using it as a benchmark tool as well for like if we see change. And then also perhaps to use as an education tool for the client. | “So saying like, look at all these areas of strength you have that you're using to support your communication. Let's be thinking about how the tools we use might map onto your incredible pragmatics and your storytelling. So let's, you know, further develop your photos and maybe attach some, you know, scripting to help you have your message come out more clearly, right. So that would probably be where I would ultimately land.” | “Yeah. I wonder. If I tend to not, I have challenges with like data sheet use, right or I, I don't tend to have a lot of papers because part of what fuels…so I guess one other possibility your thought is if it's already built into the EMR, you know that's a much more likely scenario or if it's.”  “I could very much see myself … pulling it up and referencing and like using it as a benchmark tool.” |
| 2 | “I like [the] terms of the measure.” |  |  |
| 3 |  | “I think what I would probably do is after I used it, I could use like the columns of like consistent strengths, intermittent strings, limited strengths. In my conversation with the person like so, I noticed that during this conversation consistently you. Uh, had like you seem to comprehend sentences really well.”  “Now I feel like when I listen to another video, I will be like, my mind will be like, ohh, that was a strength that they just said about themselves” |  |
| 4 | “I could totally use this in my clinical practice.”  “I think tools like that … are really great to kind of give a technical and systematic profile. For people to think and get kind of creative in that moment, they're not having to take that. You know, there's like kind of this empty step between like, OK, this is going wrong. Like this is what we're going to do about it. But like this is what's going well, this is how we can use it. This is what’s present here, which I love. So I think from like a teaching and a clinical use perspective, it is so deeply needed. Yeah, Very cool. Very cool work.” | “I especially like from the get like having your thinking set up or not you know what are the deficits where the difficult areas but. Like what, what can we actually do about it? And I think making that gap is a lot of that where that like clinical, like kind of creativity or like, you know, the magic in therapy happens.” |  |
| 5 | “I feel like a lot of these items are the stuff that I specifically comment on, like when I'm doing a new evaluation. But yeah, as a clinician, I would absolutely like this. Absolutely. I think this is more effective. But I feel like especially once I've done this a few times and kind of knew and was a little bit more familiar with each of these items, I feel like I'm really confident I could absolutely do this. After having, you know, doing an evaluation with someone or having a couple of conversations with them, record, and fill it out.” | “Not a single item on here that I think is not relevant, quite frankly. I think all of these have a lot of relevance and I could see for lots of different people certain items that these really are capturing like specific things that could be really functional goals to work on for them if they wanted to. Um, but I do also think it's nice because it captures kind of a wide variety of communication so that you can really showcase like what other areas where they are really strong and when they're doing a really fantastic job rather than stay independent.” | “I've been fortunate to, in several settings, be a research clinician, but in my current setting I'm more of a standard outpatient clinician, so I recognize that recording speech samples is typically not the norm. But like, in an ideal world, I would love to have recorded samples to at least give it a second listen, I think to comment on a few of these things.” |
| 6 | “Absolutely would use it. The scale anchors a conversation with a patient and can then also help me write my report. It gives me the terminology I need to talk about what they see, what I see, but might not have the words or tests for.” |  |  |
| 7 | “But yeah, I'm just making sure that what we're working on is really relevant to her. And I think from this 5 minute video I can get a sense of what's important to her” |  |  |
| 8 |  | “Well, it's helping me think about things in a little bit of a different light instead of deficit based, which is usually what we're thinking about thinking. About, well, what can this person do? Well, what's a strength and what's not a strength? And then not feeling the pressure to have to make a decision about someone's strengths when you didn't observe it. That I really like it. That's not always an option for some of these standardized tests, you know, or you know, like if if there's a task that the person just cannot do and. I don't know, I guess it's just kind of a little bit more functional and like like a little bit more. Like it's a little bit more realistic in terms of like what we might. Take away from like, I don't know, I'm just having a conversation with someone. How easy is it to understand what they're saying? Like how comfortable do they seem? Like do they, do we feel like we're on the same page? Are we having breakdowns? Like I feel like it leans a little bit more towards like what we are going for in terms of like real life and like real world conversation versus looking at like, you know, lexical retrieval and…which is fine too, but you know, like this seems like a lot more holistic… maybe, maybe that's the right word that I'm looking for.” |  |
| 9 | “This is really cool. I like it. I mean, I think it's like, very short, like it's short. And that I think, I think what works well is that it's kind of this structured perceptual judgment, right... I think it's super implementable.” |  | “And that it's, I mean, once you get familiar with it, you wouldn't need obviously need to be looking at it and reading each each thing and you'd start to just take notes on those things. Like I just didn't even think really about like what I should be looking for and discourse or something like that, right. But now that I have a better idea, even just... using it one time like to think about some of those pieces more right that they would be things you just think about while you're talking to your patient.” |
| 10 | “The tools support our assessment workflow. So I would definitely use the PACT and I think there it's a two-part answer as an SLP that works primarily with PPA. I think it's really beneficial in thinking about those overall aspects of language and a lot of these questions are also, you know, thinking about functional communication… so I think it's really, really important and will be helpful when assessing a patient for the first time. And secondly, I think that there this could also be used for other non SLP healthcare professionals, including neurologists. | “I think that oftentimes with a nonfluent patient you focus on the relative weakness. So in this case it might be motor speech or fluency speech fluency but doing the PACT was really helpful in highlighting [this patient’s] strengths because her discourse for instance, and her pragmatic social pragmatics and even word finding were so strong.”  “I say this because I only work with PPA for the last however many years but I think clinicians maybe focus more on word finding difficulties and compensatory strategies or even circumlocutions in the context of lvPPA and svPPA. But it's really good to be cognizant of that and you know think of that whether there is that that word finding or retrieval is a relative strength or not, so it was a good reminder as I was going through that list to think more about their or her overall lexical semantic abilities.” | “Because you know, clinic is already always at overtime, there are time constraints already in clinical settings and all you do is an MMSE for a cognitive test. And most of the things you do is conversation, whether it's getting history that's a bit more like what was the [PACT’s] first prompt. Or, you know, at having them kind of talk about their challenges and assessing their self-awareness, things like that I think already happens in clinical settings. So it could be widely applied to neurologists or other PCP's even. Because obviously the key here is to have early detection and to refer them to specialist centers as soon as possible for interventions so I think it can be applied to both settings SLP and non SLP healthcare professionals, or should be!” |
| 11 | “It does like, [feel like a screening tool] in a fantastic way too that I don't think without filling that out I, I don't think I would have walked away with those same ideas necessarily.”  “So I definitely feel like it was like comprehensive, but like fairly quick and got the wheels spinning in a way that right I don't think I necessarily would have thought otherwise”  “The four main domains I thought that there wasn't anything that I clearly felt like was missed.” | “Like I just sort of felt like, OK, this, this patient actually had quite a lot of strengths. I felt like because she, I was able to mostly get across what I believe she was trying to convey.”  “I do feel like after filling it out…I'd probably be able to walk away with like just an overall summary from that of what I felt like it was the theme of what I would address for her and it was probably. Some of that like specificity and maybe like just a little more depth on on conversation or trying to share more information other than just the kind of…surface.” | I think my first main thought is that, yeah, for the first time filling it out, it was interesting to try to, you know, when having a Likert scale like figure out what my anchors were within my own like mental sort of schema, um. Yeah, almost like, am I comparing this patient to all patients? Am I just thinking about this patient compared to themselves? You know, that that type of thing I think was hard at first to try to figure out. Ultimately, I feel like I sort of did. Uh, a combination of both. but but yes, I felt like I was able to kind of come up with a bit of a complete picture. Yeah, like a, a, a good take away. |
| 12 | “I feel like definitely clinically. And like I feel like in research, depending on like if it aligned with whatever, if it was aligning with, if I was looking at something, I was looking at language across all of these different aspects with the goal of approving speech, language pragmatics, discourse like generally. In the means like most definitely and, but definitely clinically, especially if uh, like I can imagine some of our like geriatric clinics. Like where people might come in more than once with like progressive, most of them are coming in with like some not always language related, but like like not always PPA, but you know, as a way just to track their communication could be useful.” |  |  |
| 13 | “I am really, really interested and excited actually to use it. I think it's very. It's very, it's, it's much more intuitive than you know, going through it. It's been really helpful having you sort of go through the process. I looked at the, um, the domains before and I sort of wrote more down to, to kind of familiarize myself, um.  “I’m very keen to you know to give it you know try it more when we get to that point.” | “But I felt it was it was a really good way of. Pulling out, um, aspects of someone's communication presentation, which would be really useful for therapy, you know, it's a…really good way of drilling down to the things which matter most and which have most impact on communication effectiveness?” | “I think. I think having the video is really necessary because there were certain things that, you know, I thought, Ohh, I don't know whether there was an example of that. Um, and I'd want to sort of go back and, and check, um, I think. I would struggle to do it in real time. You know, to sort of. Go between, you know, the different because you're, you're obviously, um, you know, taking in all these things on all these levels and you can't necessarily zoom into different aspects because it's all coming at you in one go.” |
