## Supplementary Table 4 for "The Progressive Aphasia Communication Toolkit (PACT): A Strengths-Based Approach to Multidomain Evaluation for Intervention"

**Supplementary Table 4. Speech-language pathologist rater agreement on the measurability and strengths-ratings of the PACT items by scale, care partner presence, and diagnosis.** Measurability was defined as the ability to evaluate a skill based on the PACT protocol and video content (0 = not measurable, 1 = measurable). Strength agreement was defined on a binary to see if raters agreed on whether a skill was a strength (not a strength = not a strength *or* limited strength, strength = consistent strength *or* intermittent strength) to capture overarching consistency in strength ratings.

| **Factor** | **Scale** | **Care Partner** | **Diagnosis** | **Rater Agreement** |
| --- | --- | --- | --- | --- |
| **Measurability** | Speech and Voice |  |  | 0.996 |
|  | Language |  |  | 0.892 |
|  | Social-Pragmatics |  |  | 0.918 |
|  | Discourse |  |  | 0.941 |
|  |  | Present |  | 0.918 |
|  |  | Not Present |  | 0.911 |
|  |  |  | svPPA | 0.961 |
|  |  |  | nfvPPA | 0.919 |
|  |  |  | lvPPA | 0.903 |
|  |  |  | Mixed | 0.875 |
|  | Speech and Voice |  | svPPA | 1.000 |
|  |  |  | nfvPPA | 1.000 |
|  |  |  | lvPPA | 1.000 |
|  |  |  | Mixed | 0.984 |
|  | Language |  | svPPA | 1.000 |
|  |  |  | nfvPPA | 0.861 |
|  |  |  | lvPPA | 0.844 |
|  |  |  | Mixed | 0.861 |
|  | Social-Pragmatics |  | svPPA | 0.900 |
|  |  |  | nfvPPA | 0.875 |
|  |  |  | lvPPA | 0.850 |
|  |  |  | Mixed | 0.734 |
|  | Discourse |  | svPPA | 0.933 |
|  |  |  | nfvPPA | 0.958 |
|  |  |  | lvPPA | 0.933 |
|  |  |  | Mixed | 0.938 |
| **Strength** | Speech and Voice |  |  | 0.849 |
|  | Language |  |  | 0.853 |
|  | Social-Pragmatics |  |  | 0.835 |
|  | Discourse |  |  | 0.730 |
|  |  | Present |  | 0.747 |
|  |  | Not Present |  | 0.888 |
|  |  |  | svPPA | 0.683 |
|  |  |  | nfvPPA | 0.886 |
|  |  |  | lvPPA | 0.890 |
|  |  |  | Mixed | 0.809 |
|  | Speech and Voice |  | svPPA | 0.975 |
|  |  |  | nfvPPA | 0.594 |
|  |  |  | lvPPA | 0.925 |
|  |  |  | Mixed | 0.902 |
|  | Language |  | svPPA | 0.975 |
|  |  |  | nfvPPA | 0.986 |
|  |  |  | lvPPA | 0.822 |
|  |  |  | Mixed | 0.958 |
|  | Social-Pragmatics |  | svPPA | 0.625 |
|  |  |  | nfvPPA | 1.000 |
|  |  |  | lvPPA | 1.000 |
|  |  |  | Mixed | 0.663 |
|  | Discourse |  | svPPA | 0.467 |
|  |  |  | nfvPPA | 0.972 |
|  |  |  | lvPPA | 0.800 |
|  |  |  | Mixed | 0.682 |

*svPPA: semantic variant primary progressive aphasia*

*nfvPPA: nonfluent variant primary progressive aphasia*

*Mixed: mixed presentation of primary progressive aphasia*

*lvPPA: logopenic variant primary progressive aphasia*
